# Calendar Period Estimation of Probabilities of Transition in Drug Development

**DOI:** 10.1101/2025.05.15.25327693

**Authors:** Ernesto Schirmacher, Clifton Chow, Fred Ledley

## Abstract

This paper examines the intricate and high-risk process of drug development. A compound’s journey from target validation to market launch can span 10 to 15 years, with an average cost of one to two billion USD per successful drug. Despite extensive pre-clinical work, only about 10% of drug candidates entering human clinical trials achieve US Federal Drug Administration approval. The primary causes of failure during the three phases of clinical trials include lack of clinical efficacy, unmanageable toxicity, poor drug-like properties, and lack of commercial needs or strategic planning.

The study aims to measure the transition probability of drugs from Phase I to clinical approval and offers a broad comparison between drugs with different characteristics (therapeutic indication, mechanism of action, target, and modality). We employ a large industry database, Pharmaprojects, which tracks the development of drug candidates globally from pre-clinical stages through market launch and withdrawal or discontinuation (if it occurs). Thus, it provides a ‘complete’ picture of both successful and unsuccessful drugs, albeit limited to publicly available information.

Nearly all prior work on the estimation of transition probabilities has used a traditional longitudinal design. We take a cross-sectional approach which allows for a more dynamic understanding of the process as it unfolds over time and offers the possibility of detecting and measuring the calendar year impacts of policy and practice changes in a timely manner.

We construct life tables for each phase of clinical trials between 2002 and 2022 to estimate the raw hazard of transition and graduate them with a generalized additive model to produce a smooth hazard and corresponding conditional probabilities of transition and the overall probability of success.

Our analysis shows that the propensity to transition out of each of the three phases of clinical trials behaves differently, changes over time, and is heavily influenced by certain drug characteristics, such as, the therapeutic indication or the mechanism of action. Moreover, we find positive trends in the overall probability of success for certain classes of drugs suggesting that the industry is improving its productivity.

**Practical Applications Summary:** With our results we can estimate various conditional transition probabilities given that a drug has been under development for some time and combine them to provide probabilistic results for a portfolio of drugs.

## 1. Introduction

The discovery and development of drugs for human use is a complex, time consuming, expensive, and very risky endeavor. The entire process from target validation to market launch can take 10 to 15 years and the average cost per successful drug can be between one and two billion USD (Kramer, Sagartz, and Morris 2007; DiMasi, Grabowski, and Hansen 2016). The discovery process can be split into the following components: target validation (*≈* 1.5 years), compound screening (*≈* 1.5 years), lead optimization (*≈* 1.5 years), pre-clinical tests (*≈* 1 year), three phases of human clinical trials (Phase I *≈* 1.5 years, Phase II (*≈* 2.5 years), and Phase III (*≈* 2.5 years), and approval to launch (*≈* 1.5 years) (Sun et al. 2022).

Despite all the pre-clinical work done (approximately 5 years of research and development) to come up with a new drug candidate that is ready for human clinical trials, only about 10% of these candidates will eventually be approved for human use (Dowden and Munro 2019; Sun et al. 2022). The reasons for failure during the clinical trials are varied, but can be broken down into four main categories (Sun et al. 2022):

1. lack of clinical efficacy (40%–50%)
2. unmanageable toxicity (30%)
3. poor drug-like properties (10%–15%)
4. lack of commercial needs and poor strategic planning (10%)

We will not delve into the reasons for failure. Rather we are interested in measuring the conditional probability of transition out of a clinical trial phase given the amount of time a drug has spent in a particular phase of development and how certain broad characteristics of the drug affect these probabilities and consequently the total time spent in a clinical trial.

The Phase I trials are mainly concerned with the safety of a drug. A small number of participants (usually between 20 and 80) are given a small dose of the drug and are followed up carefully to understand any side effects and how the human body and the drug interact (pharmacokinetics and pharmacodynamics). The dosage is increased until side effects or toxicity is evident to determine the largest dose that would be safe to use. Phase II trials investigate whether or not the drug works as intended and to uncover possible rare side effects. These trials usually enroll between a few dozen and 300 people. Phase III trials are larger (between a few hundred and 3,000 individuals) and the goal is to determine if the new drug is better and/or safer than the current standard treatment.

If a new drug successfully completes these three phases of clinical trials, then, in the United States, the drug’s sponsor can apply for approval with the Food and Drug Administration (FDA). The FDA reviews all the information from the clinical trials and decides whether or not to approve the drug for use in patients with the same illness as those who were tested. The use of an approved drug for a different illness requires a new set of human clinical trials to gather the appropriate evidence of its safety and efficacy in treating the new illness, and a new application for approval with the FDA.

Past studies, such as (Hay et al. 2014; DiMasi, Grabowski, and Hansen 2016; Yamaguchi, Kaneko, and Narukawa 2021), have investigated the time spent in the different phases of human clinical trials and/or quantified the probability of transitioning out of each phase and the overall probability that a drug candidate will eventually be approved for human use. These studies have taken a cohort of drugs and followed their experience over a number of years and provided an estimate of the probability of success for each phase and for the entire experience period. Table 1 displays the estimates obtained for the overall probability of success (from Phase I to approval) for the drugs studied.

**Table 1.** Estimates of the overall probability of success from Phase I clinical trials to approval for use in humans for different studies.

| Source | Publication<br>Year | Study<br>Period | Probability of<br>Success (in %) |
| --- | --- | --- | --- |
| Hay et al. | 2014 | 2003–2011 | 10.4 |
| DiMasi, Grabowski, and Hansen | 2016 | 1995–2007 | 11.8 |
| Yamaguchi, Kaneko, and Narukawa | 2021 | 2000–2010 | 12.8 |

This approach requires observing the drugs in the cohort for a long period of time in order to reliably estimate the proportion that succeed. Note that the publication dates for two of the three sources listed in Table 1 are about a decade after the end date of the cohort of drugs that are part of the investigation. Such a long delay is not ideal if we are trying to understand some of the forces that may influence the speed at which drugs navigate the three phases of human clinical trials. In particular, it would be hard to detect and measure the impact of policy & practice changes in a timely manner.

Our approach focuses on estimating the conditional probabilities of transition given the amount of time under development and are based on a calendar period of observation. We take a cross-sectional view of the data as opposed to a longitudinal view (as in the references in Table 1). Hence, we are estimating a series of *current*transition tables as calendar time progresses. The rest of the paper is organized as follows: the next section discusses the data and its characteristics. Section 3 describes the methods used to estimate transition probabilities, their smoothing, and how various characteristics of a drug affect them. Section 4 shows an application of our methods to the evaluation of a sample portfolio of drug. In the last section we provide some concluding remarks.

## 2. Data Characteristics

The data used in this study comes from Pharmaprojects (Citeline 2003); a global database that tracks the research and development of drug candidates, intended for use on humans, around the world. Data collection began in 1980 and contains information from all phases of the drug development process (pre-clinical, human trials, approvals, discontinuation, and/or removal from the market). Regardless of the fate of a drug candidate, the information is recorded and retained in the database. Hence, we have a ‘complete’ picture of both successful and unsuccessful drugs. Pharmaprojects uses information in the public domain to update its records and thus it is limited to what drug sponsors are willing to share with the public.

The Pharmaprojects database, as of May 2023, contains a total of 92,308 compounds excluding vaccines. The database covers all diseases, indications, and therapeutic areas as well as mechanisms of action, biological targets, chemical properties, origin, and delivery routes plus key events, such as, the starting dates of clinical trials, the approval dates of a drug, and their market launch. It is important to note that the ending dates for clinical trials are not recorded in the database. Moreover, we have noticed that many dates have been recorded in the middle of the month and so we have made the assumption that dates are only accurate to the month. Hence, our calculations only use year and month.

In our study, we will estimate the conditional probabilities of transition out of a phase given how long a drug has been in development during this phase. Unfortunately, many of these drugs do not have information regarding the starting dates of Phase I, II, and III clinical trials or their approval date. Moreover, because the database tracks the events of a drug globally, we may have multiple approval dates (different countries) as well as multiple starting dates for each of the clinical trial phases (multiple trials in one country or multiple countries with trials). For each drug and each clinical phase or approval, we have selected the earliest available dates. Table 2 shows the number of drugs that have zero, one, two, three, or four dates (starting Phase I, II, III or approval) available. Most of the drugs (nearly 70,000) that have no clinical trial dates. They can be categorized as follows: no development reported (*n* = 46, 959), in a pre-clinical stage (*n* = 11, 111), have been discontinued (*n* = 8, 824), have been launched (*n* = 2, 315), or any of another eight categories none of which has more than 100 drugs in them (*n* = 389). Removing all drugs that do not have at least one clinical trial starting phase date leaves us with 22,710 drugs to work with.

**Table 2.** Number of drugs categorized by the number of dates available for Phase I, II, III, and Approval.

| #Dates | #Drugs |
| --- | --- |
| Zero | 69,598 |
| One | 15,464 |
| Two | 5,269 |
| Three | 1,510 |
| Four | 467 |
| Total | 92,308 |

This subset of drugs have starting dates that span the period from 1989 to 2023. Figure 1 shows the number of drugs starting a clinical trial by phase from 1990 to 2022 (omitting the partial year of data at both ends). There is steady increase through the 1990’s and 2000’s, but after 2012 the trends increase sharply for all three phases.

**Figure 1.**
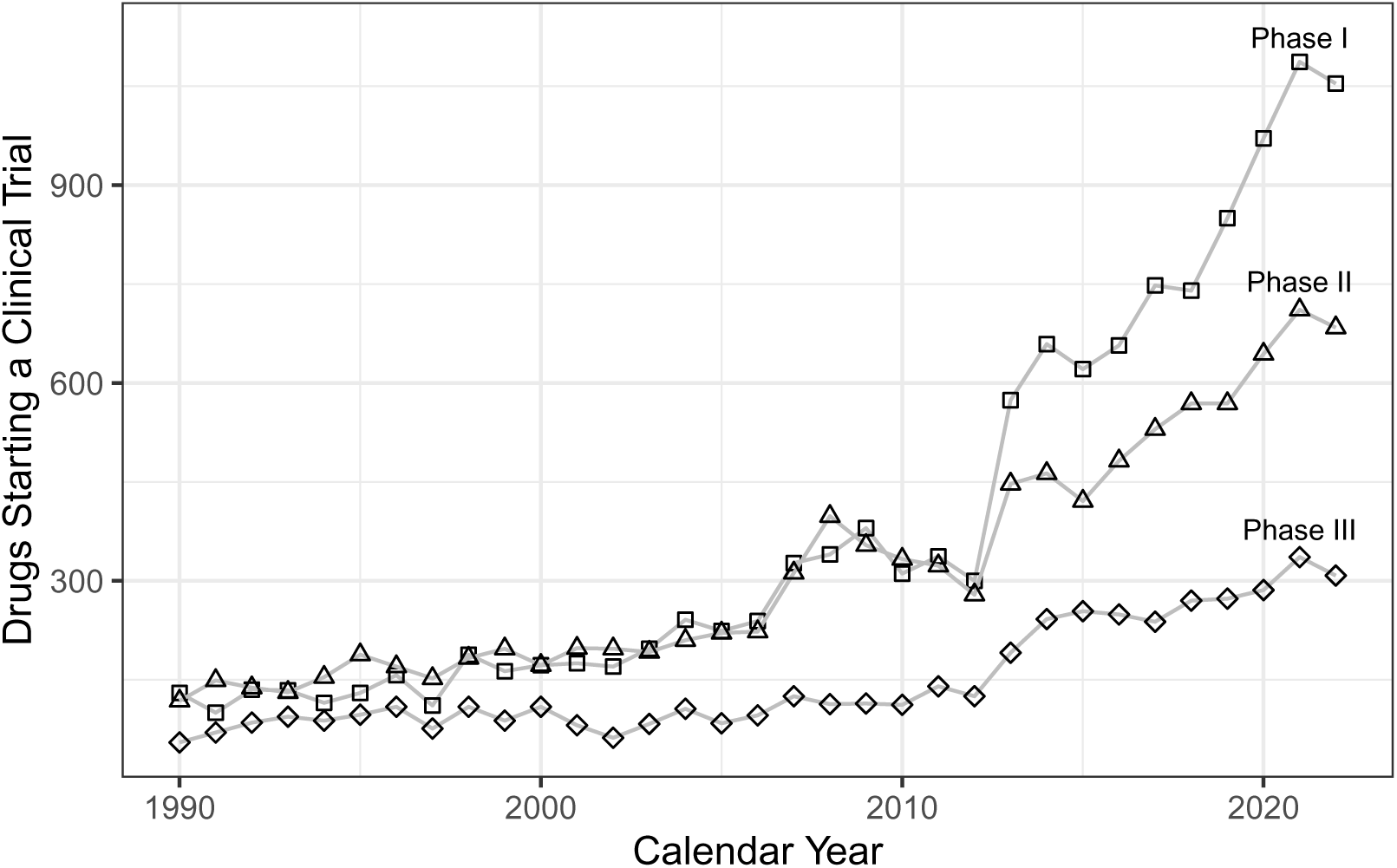
Number of drugs starting a clinical trial by phase and calendar year (omitting partial years 1989 and 2023).

As an example of the key events and dates information we have available for each drug consider **ripretinib**; a cancer medication that has been developed to treat advanced gastrointestinal stromal tumors in adults. Table 3 shows a partial list of the information available in Pharmaprojects. Notice that we have several filing dates (in the United States and Australia), several approval dates (United States and Canada), and no starting date for Phase II clinical trials. Since Pharmaprojects does not track the end date of clinical trials, we use the difference in two starting dates as a proxy for the amount of time spent in a particular phase of development. For ripretinib, the amount of time spent in Phase I is 2 years and 3 months and in Phase III together with the time necessary to file and receive approval would be 2 years and 4 months.

**Table 3.**
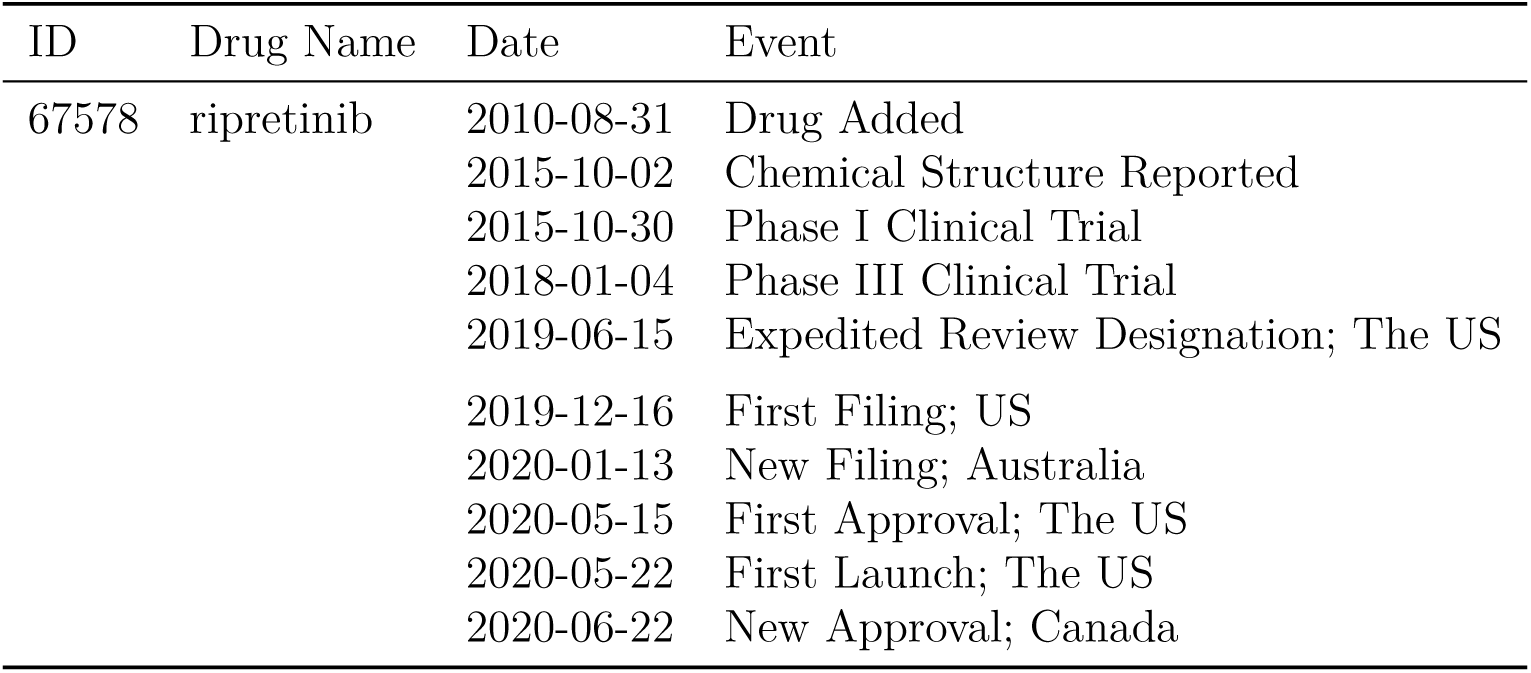
Partial list of the event information available for ripretinib (ID 67578). This drug, like most in the database, has some missing information. In this case, we do not have a Phase II clinical trial date.

Most of the drugs in the database, like ripretinib, have some dates for clinical trials and/or approval missing. In cases where we have a starting date but the next starting date is missing, we use the next available date to measure the time in this phase. For example, we have a Phase I starting date but the Phase II starting date is missing. Then we would look for the Phase III or approval dates and use the earliest to compute the time spent in Phase I. If neither a Phase III or Approval date is available, then we have a censored observation.

Table 4 shows the number of drugs, the median and mean time spent in a clinical trial phase split by those drugs that transitioned successfully and those that have been censored. The time in phase estimates for successful drugs (first row in Table 4) is a lower bound on the actual time spent in a phase since these estimates do not incorporate any information from the censored group. From this table, we can see that the overall time spent in human clinical trials to bring a drug to market is at least about 10 years. Figure 2 shows the mean and median times to transition for successful drugs by phase and by calendar period of observation. The data has been grouped in 2-year periods by starting phase date with the first group 1991-1992 and the last group 2015-2016. We are omitting calendar years 2017 to 2023 because these years would not have enough time to develop and so their estimates would be biased downwards.

**Figure 2.**
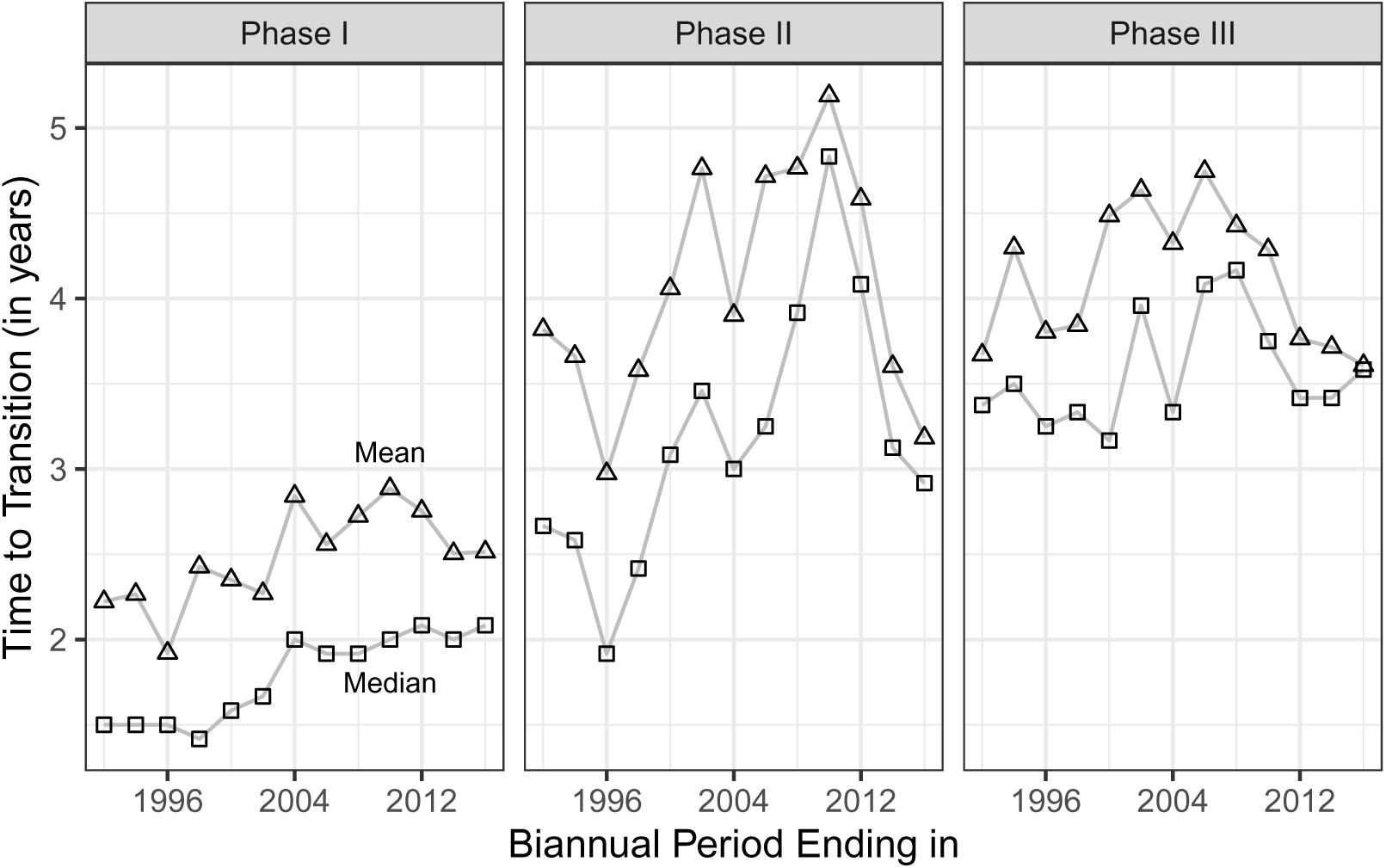
Mean and median time to transition (in years) for drugs that have transitioned successfully. Data has been grouped biannually ending from 1992 to 2016.

**Table 4.**
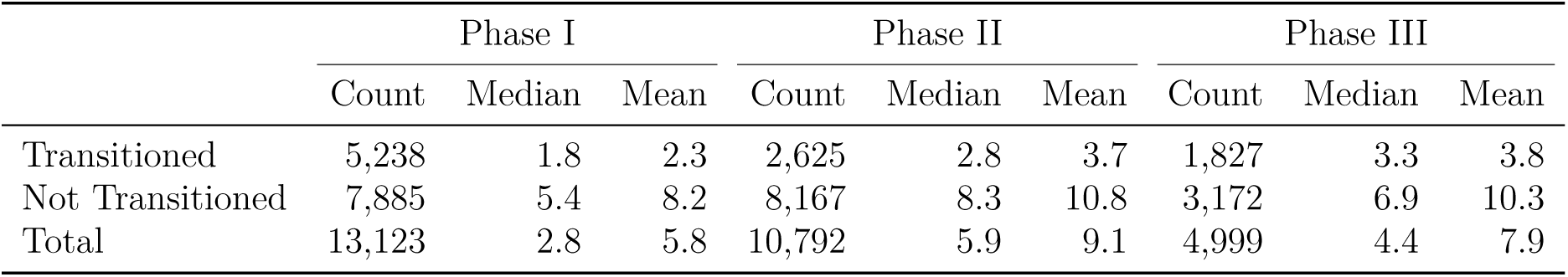
Number of drugs along with median and mean time (in years) by phase to either transition out of phase successfully or be censored for all drugs under consideration.

|  | Phase I |  |  | Phase II |  |  | Phase III |  |  |
| --- | --- | --- | --- | --- | --- | --- | --- | --- | --- |
|  | Count | Median | Mean | Count | Median | Mean | Count | Median | Mean |
| Transitioned | 5,238 | 1.8 | 2.3 | 2,625 | 2.8 | 3.7 | 1,827 | 3.3 | 3.8 |
| Not Transitioned | 7,885 | 5.4 | 8.2 | 8,167 | 8.3 | 10.8 | 3,172 | 6.9 | 10.3 |
| Total | 13,123 | 2.8 | 5.8 | 10,792 | 5.9 | 9.1 | 4,999 | 4.4 | 7.9 |

Note that the raw times to transition displayed for Phase I in Figure 2 have been increasing. A similar increasing trend is evident for Phase II up until 2010. Thereafter, we see a strongly decreasing pattern. For Phase III, we also have an increasing pattern up until around 2006 or 2008 and then a decreasing one.

In addition to starting dates for clinical trials and drug approval dates, Pharmaprojects has additional classification variables; such as, *modality*, *mechanism of action*, *target*, and *indication*. The first three variables have mutually exclusive categories, but indication does not. Many drugs have multiple indications and the top five categories are: Anti-cancer (*n* = 6, 567), Neurological (*n* = 4, 471), Alimentary/Metabolic (*n* = 3, 431), Cardiovascular (*n* = 2, 450), and Anti-infective (*n* = 2, 448). The remaining categories (musculoskeletal, unspecified, genitourinary (including sex hormones), rare diseases, blood and clotting, dermatological, immunological, miscellaneous, hormonal (excluding sex hormones), sensory, and anti-parasitic) comprise about 35% of the total.

Pharmaprojects provides four categories for modality: Chemical (*n* = 15, 342), Biologic (excluding mAb) (*n* = 5, 278), Monoclonal Antibody (mAb) (*n* = 1, 291), and Plant (*n* = 799). For mechanism of action there are six categories: Inhibitor (*n* = 8, 118), Agonist (*n* = 3, 542), Antagonist (*n* = 3, 733), Stimulant (*n* = 2, 058), Other (*n* = 1, 687), and Unidentified (*n* = 3, 572). There are seven target categories: Unspecified (*n* = 6, 721), Receptor (*n* = 6, 537), Other (*n* = 5, 683), Enzyme (*n* = 3, 316), Ion Channel (*n* = 387), Ligand (*n* = 59), and Transporter (*n* = 7).

## 3. Methods and Results

Over the last two decades there have been many studies (DiMasi, Hansen, and Grabowski 2003; Abrantes-Metz, Adams, and Metz 2004; Kola and Landis 2004; Hay et al. 2014; DiMasi, Grabowski, and Hansen 2016; Smietana, Siatkowski, and Møller 2016; Wong, Siah, and Lo 2019; Yamaguchi, Kaneko, and Narukawa 2021) quantifying the probability of success for each individual phase of the human clinical trials as well as the overall probability of approval for candidate drugs. All of these papers have looked longitudinally at a cohort of drugs. Some of these (DiMasi, Hansen, and Grabowski 2003; DiMasi, Grabowski, and Hansen 2016) are based on bespoke surveys of pharmaceutical companies while others (Abrantes-Metz, Adams, and Metz 2004; Hay et al. 2014; Wong, Siah, and Lo 2019; Yamaguchi, Kaneko, and Narukawa 2021) used large industry databases.

We use a large industry database, but instead of selecting a cohort of drugs and study them longitudinally, we are interested in the probabilities of transition by calendar year periods. Hence, we take a cross-sectional slice of time, an observation period, and study the transitions that occur during this window.

Our point of departure is to summarize the transition experience of candidate drugs that were in active development during a particular period of observation. Table 5 shows the experience, organized in a life table, for Phase I drugs during the calendar years 2017, 2018, and 2019.

**Table 5.** A life table for Phase I during the experience period 2017–2019. The first column shows the amount of time a drug has been in Phase I. The second column is the exact time drugs have been at risk of transitioning. The third column shows the number of successful transitions out of this phase that occurred during the time interval.

| Time in<br>Phase | Exact<br>Exposure | Transitions | Hazard | Probability<br>of Transition |
| --- | --- | --- | --- | --- |
| [0,1) | 2,167.8 | 217 | 0.100 | 0.095 |
| [1,2) | 1,753.0 | 263 | 0.150 | 0.139 |
| [2,3) | 1,443.0 | 196 | 0.136 | 0.127 |
| [3,4) | 1,253.3 | 98 | 0.078 | 0.075 |
| [4,5) | 1,046.6 | 55 | 0.053 | 0.051 |
| [5,6) | 794.8 | 28 | 0.035 | 0.035 |
| [6,7) | 596.4 | 12 | 0.020 | 0.020 |
| [7,8) | 521.0 | 12 | 0.023 | 0.023 |
| [8,9) | 550.2 | 8 | 0.015 | 0.014 |
| [9,10) | 518.8 | 8 | 0.015 | 0.015 |
| [10,11) | 495.0 | 3 | 0.006 | 0.006 |
| [11,12) | 392.3 | 0 | 0.000 | 0.000 |
| [12,13) | 341.8 | 3 | 0.009 | 0.009 |

<sup>a</sup> Hazard = Transitions / Exact Exposure
<sup>b</sup> Probability of Transition = $1 - \exp(-\text{Hazard})$

The first column, Time in Phase, shows a time interval representing the amount of time a drug has been under development. The second column, Exact Exposure, shows the exact amount of time drugs have been exposed to the risk of transitioning during the observation period. The next column, Transitions, counts the number of drugs that have successfully transitioned out of this phase. The hazard column is the ratio of number of transitions to exact exposure. The last column, Probability of Transition, gives an estimate of the conditional probability of transition given that a drug is still in active development via the formula *q_t_* = 1 *−* exp(*−*Hazard), where *q_t_* represents the probability that a drug that has been under development for *t* years and has not yet experienced a transition will do so within the next year.

Similar tables can be constructed for Phase II and III and other observation periods. All of these tables show a similar pattern for the hazard of transition as that shown for Phase I; namely, it starts at a medium level for the time interval [0, 1) and rises sharply to a maximum over the next few time years and thereafter it steadily decreases. The hazard pattern for each phase during the experience period 2017–2019 are displayed in Figure 3, where we have added a smooth estimate of the hazard. This smooth estimate is computed by fitting a log-link Poisson generalized additive model (Hastie and Tibshirani 1999; Wood 2006) to the transition status of each drug and a linear predictor with a smooth for the time in phase and an offset equal to the logarithm of the exact exposure.

**Figure 3.**
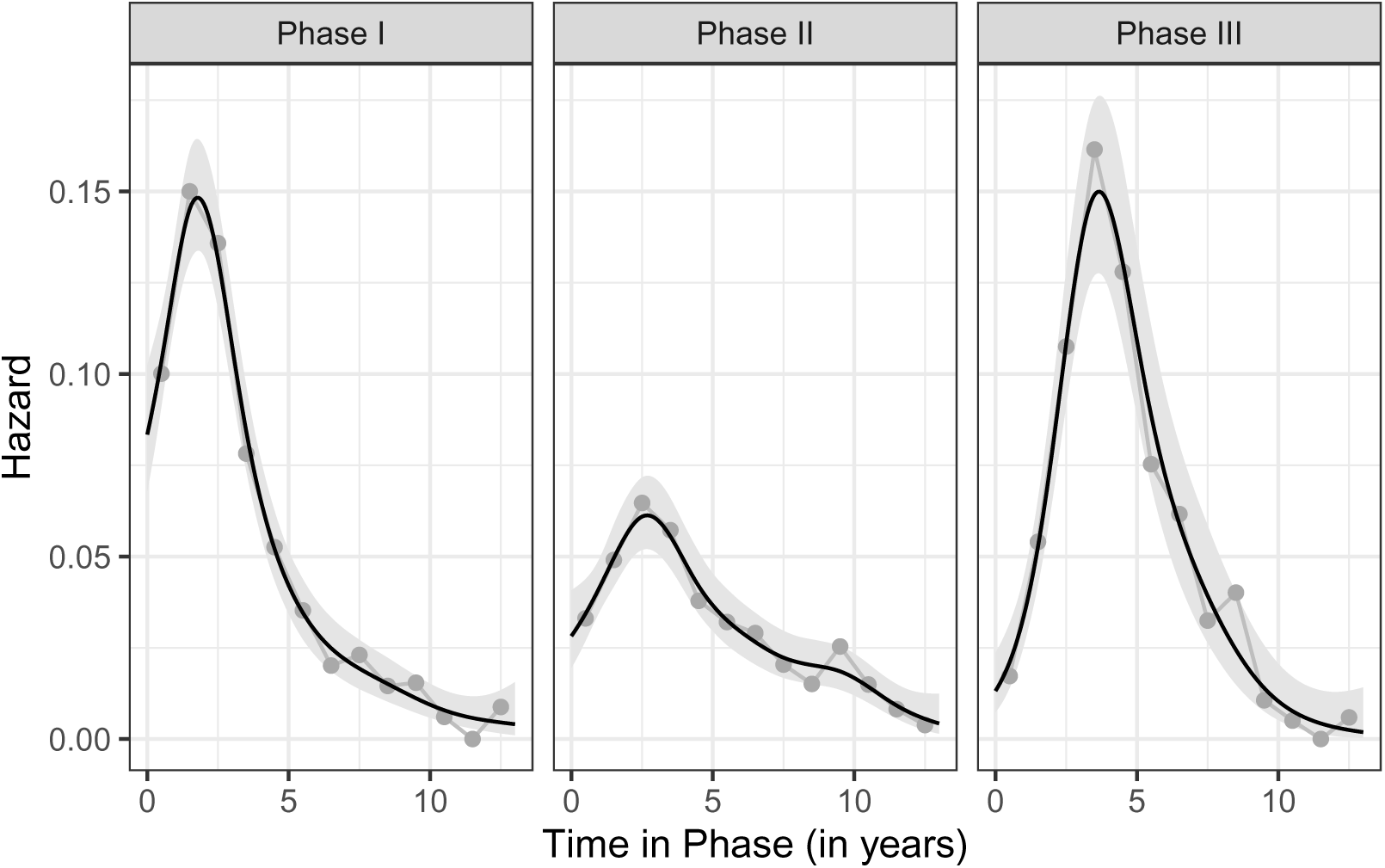
The actual hazard (points) and a smooth estimate (solid curve) of the hazard for Phase I, II, and III during the experience period 2017–2019. The shading behind the solid curve represents a pointwise 95% confidence interval for the smooth hazard.

From Figure 3 we see that the hazard for Phase II is much lower than those for Phase I or Phase III. Transitioning out of a Phase II clinical trial is a much harder than doing so out of Phase I or Phase III. Phase II trials have a much higher threshold to surpass (showing that the drug works as intended) than either Phase I (concerned with safety) or Phase III (is safer/better than current treatments).

The hazard in all three phases show a peak around 2.5 years of development work. The shape of the smooth hazard curves are reasonable. In all three phases, it takes some time for a drug to start working and its effects to be seen; hence the hazard of transitioning out of the current phase increases until it reaches a peak. Thereafter, we see a steady decrease in the hazard. Careful inspection of Phase II in the time interval [7.5, 10] reveals that the hazard plateaus before commencing a steeper decline after 10 years. One plausible scenario for such an effect might be that a drug that was initially intended for one indication did not show promise, but it may have been re-purposed for a different indication.

Based on the smooth hazard curves, we construct survival functions and cumulative incidence functions, see Figure 4, for all three phases of development and thus can estimate the overall probability of approval. For the experience period 2017–2019, the probability of transitioning out of Phase I is 47.2%, out of Phase II is 32.4%, and out of Phase III it is 50.4%; Hence, the overall probability of success is the product of these three probabilities, yielding 7.7%.

**Figure 4.**
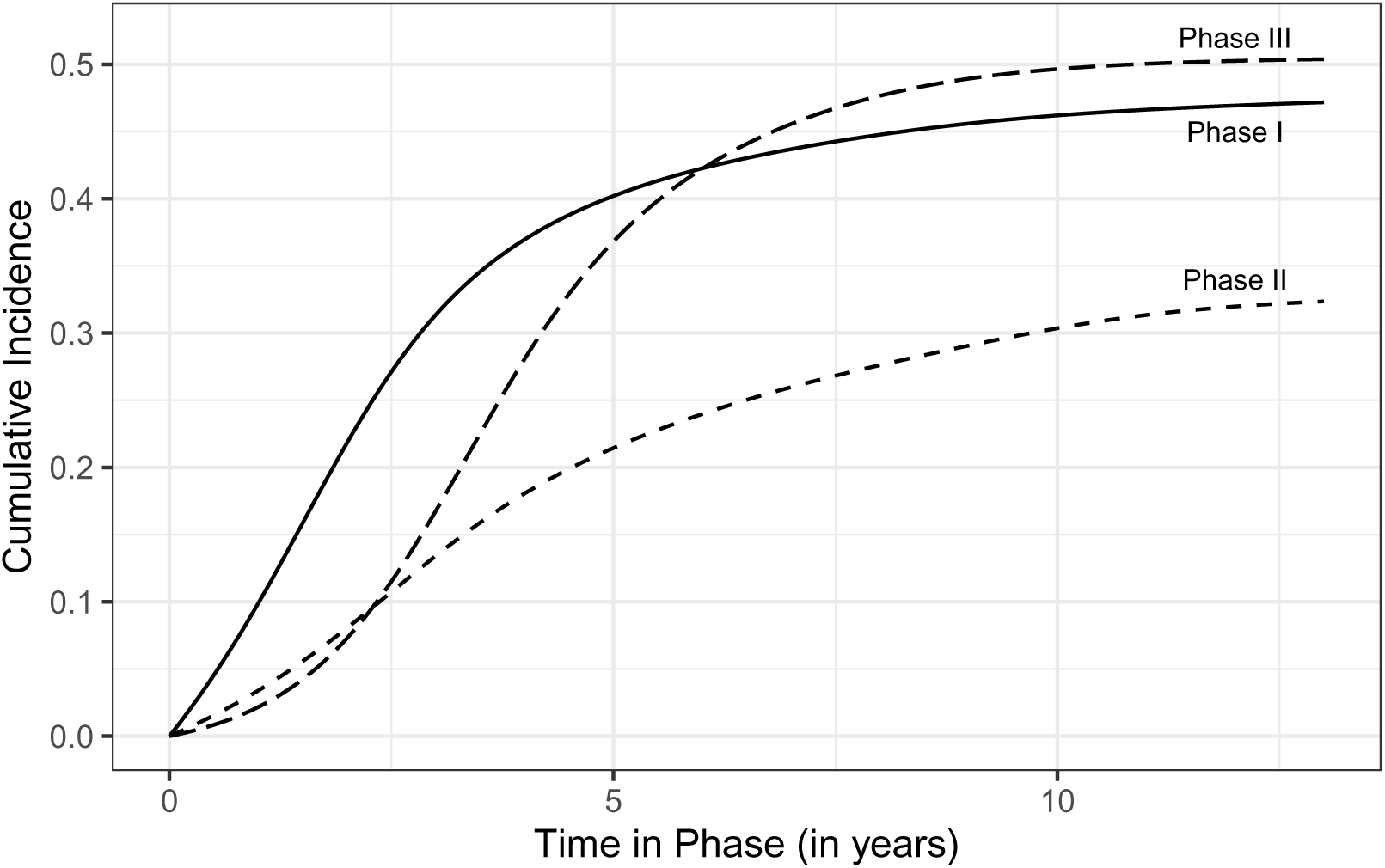
Cumulative incidence curves by clinical trial phase for the experience period 2017–2019.

Table 6 shows the probabilities of success for each phase as well as the probability of approval (the product of phase probabilities). The last two columns are a 95% confidence interval for the probability of approval and are based on a bootstrap (Efron and Tibshirani 1998) simulation with 200 replicates.

**Table 6.** Probabilities of success for all drugs by phase and experience period. The probability of approval is the product of the three phase probabilities. The 95% confidence interval (CI) is for the overall probability of approval.

| Period | All Compounds |  |  | Approval | Approval 95% CI |  |
| --- | --- | --- | --- | --- | --- | --- |
|  | Phase I | Phase II | Phase III |  | Low | High |
| 2002–2004 | 0.524 | 0.235 | 0.389 | 0.048 | 0.036 | 0.061 |
| 2005–2007 | 0.521 | 0.280 | 0.423 | 0.062 | 0.051 | 0.075 |
| 2008–2010 | 0.522 | 0.241 | 0.441 | 0.056 | 0.044 | 0.068 |
| 2011–2013 | 0.479 | 0.224 | 0.463 | 0.050 | 0.041 | 0.059 |
| 2014–2016 | 0.473 | 0.310 | 0.516 | 0.075 | 0.065 | 0.087 |
| 2017–2019 | 0.472 | 0.324 | 0.504 | 0.077 | 0.067 | 0.086 |
| 2020–2022 | 0.480 | 0.327 | 0.458 | 0.072 | 0.064 | 0.079 |

Note that the Phase I probabilities hover slightly above 52% up until 2010 and then drop to around 47% and remain stable at this level. The Phase II probabilities hover around 25% for the first four periods (until 2013) and then jump up to a new level near 32% and increase slightly over the last two periods. For Phase III, we see a strong increase in probabilities through the 3-year experience periods ending in 2016 and thereafter we have a decline. The overall probability of success, from Phase I to drug approval, is shown in the column labeled ‘Approval.’ The rates hover around 5.4% during the first four periods, and then jump up to a new level around 7.5% for the last three periods. These probabilities are shown in Figure 5.

**Figure 5.**
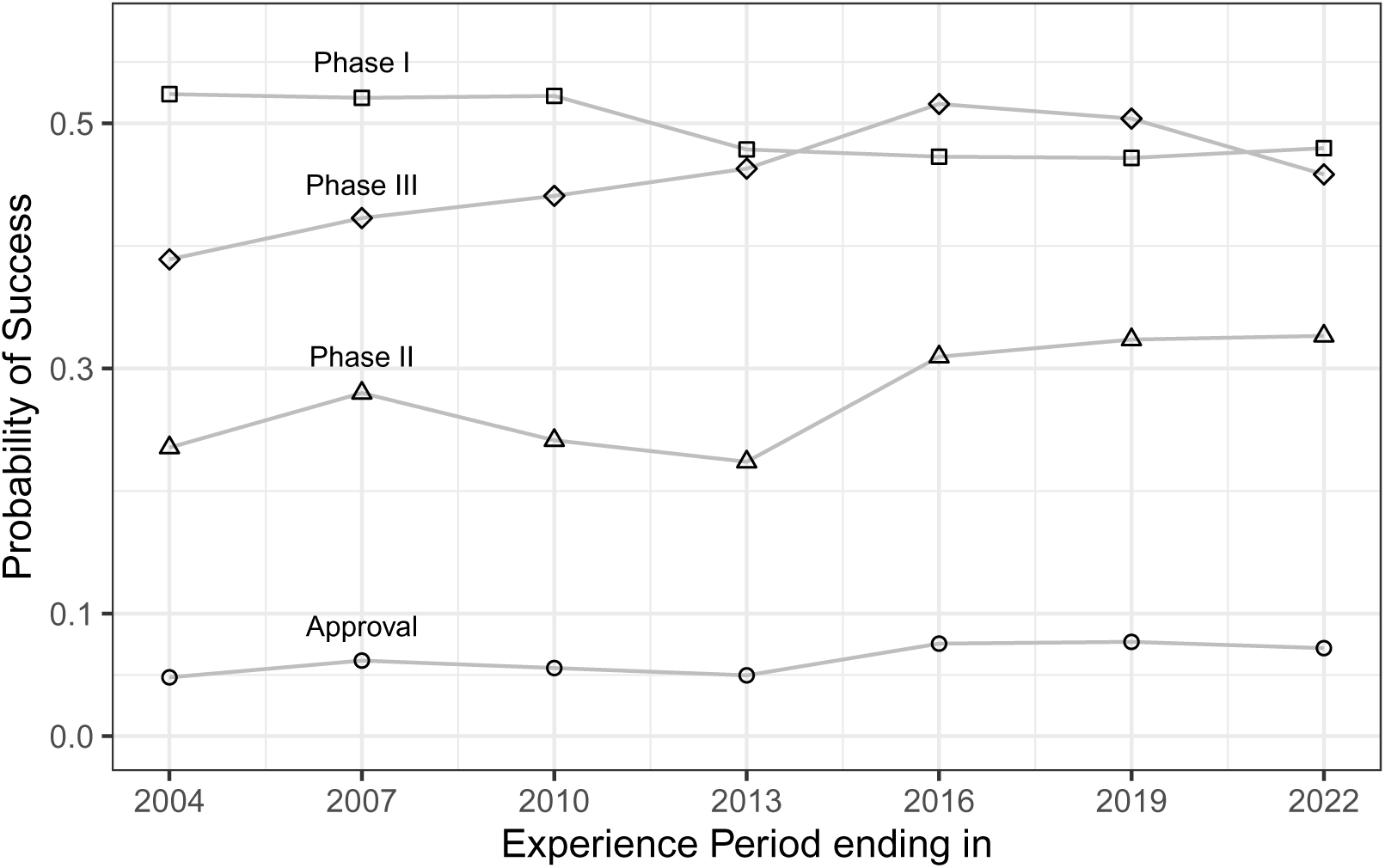
Probabilities of success for all drugs by experience period and phase. The points labelled ‘Approval’ are the product of the probabilites of success for all three phases.

These results give us an aggregate view of all the drugs under development worldwide during the last 20 years. But there are significant differences in these probabilities depending on various characteristics of the drugs involved. In the next sections we study four different characteristics: modality, target, mechanism of action, and indication.

### 3.1. Modality

Modality refers to the provenance of the drug and PharmaProjects uses four categories: chemical (*n* = 15, 342), biologic (excluding mAb) (*n* = 5, 278), monoclonal antibody (mAb) (*n* = 1, 291), and plant (*n* = 799). We have decided not to analyze the plant category.

Chemical drugs are typically manufactured from small molecules synthesized chemically from well-defined chemical structures. The manufacturing process is generally reproducible and well-established because they target specific molecules or biochemical processes. Common examples of chemical drugs in the market include aspirin and many statins that lower cholesterol. Chemical drugs are also less likely to provoke an immune response in the body and are often less expensive to manufacture when compared to other drug categories.

Monoclonal antibody (mAb) drugs are man-made proteins that mimic the way human antibodies work in the immune system. The science and engineering know-how behind these drugs is better understood and the probabilities of success for these drugs reflect this.

Biologic drugs are more complex to manufacture than other forms of pharmaceuticals. These drugs are produced from complex molecules derived from living organisms such as bacteria or yeast. They involve complex procedures that are often more intricate such as fermentation, cell culture and purification. Unlike other drugs, such as chemical drugs, that are not immunogenic, biologic drugs may trigger immune responses that can weaken their efficacy and increase adverse situations in the body. Some examples of biologics are those targeted to treat anemia, cystic fibrosis, diabetes, hepatitis, and transplant rejection. Because these drugs require more specialized technologies and are complex to produce, they are often more expensive to develop and manufacture.

For example, during the 2017–2019 experience period the hazard of transitioning out of any development phase for monoclonal antibody drugs was significantly higher compared to all other drugs as shown in Figure 6. For many of the observations periods, monoclonal antibody drugs have higher hazards than biologics (xmAb) and chemical drugs, especially during Phase I and Phase II.

**Figure 6.**
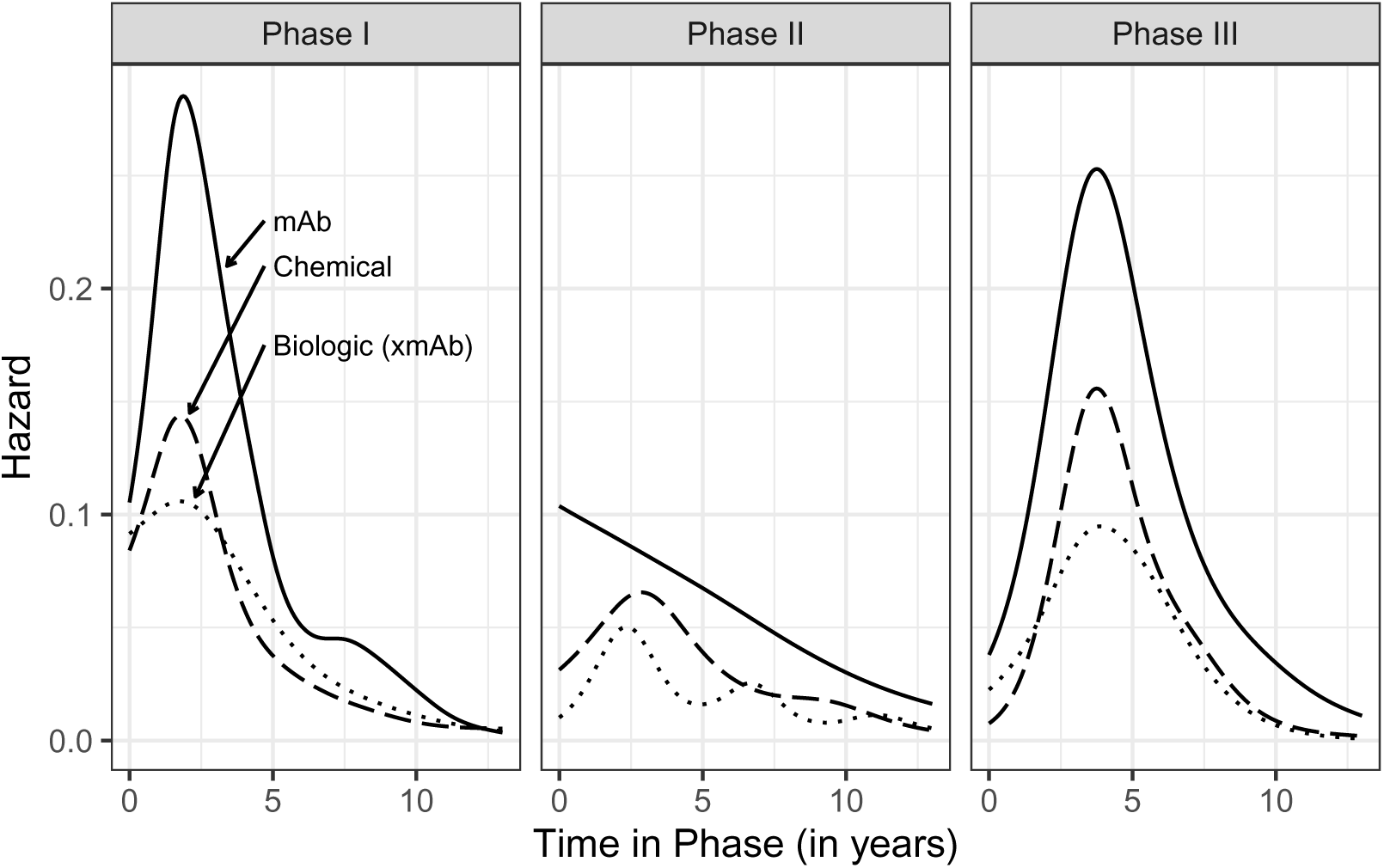
Estimated smooth hazard of transition for the experience period 2017–2019 by development phase and modality. Note that monoclonal antibody (mAb) drugs have a higher hazard of transition than all other modalities across all phases. The biologic category excludes monoclonal antibody (mAb) drugs.

**Figure 7.**
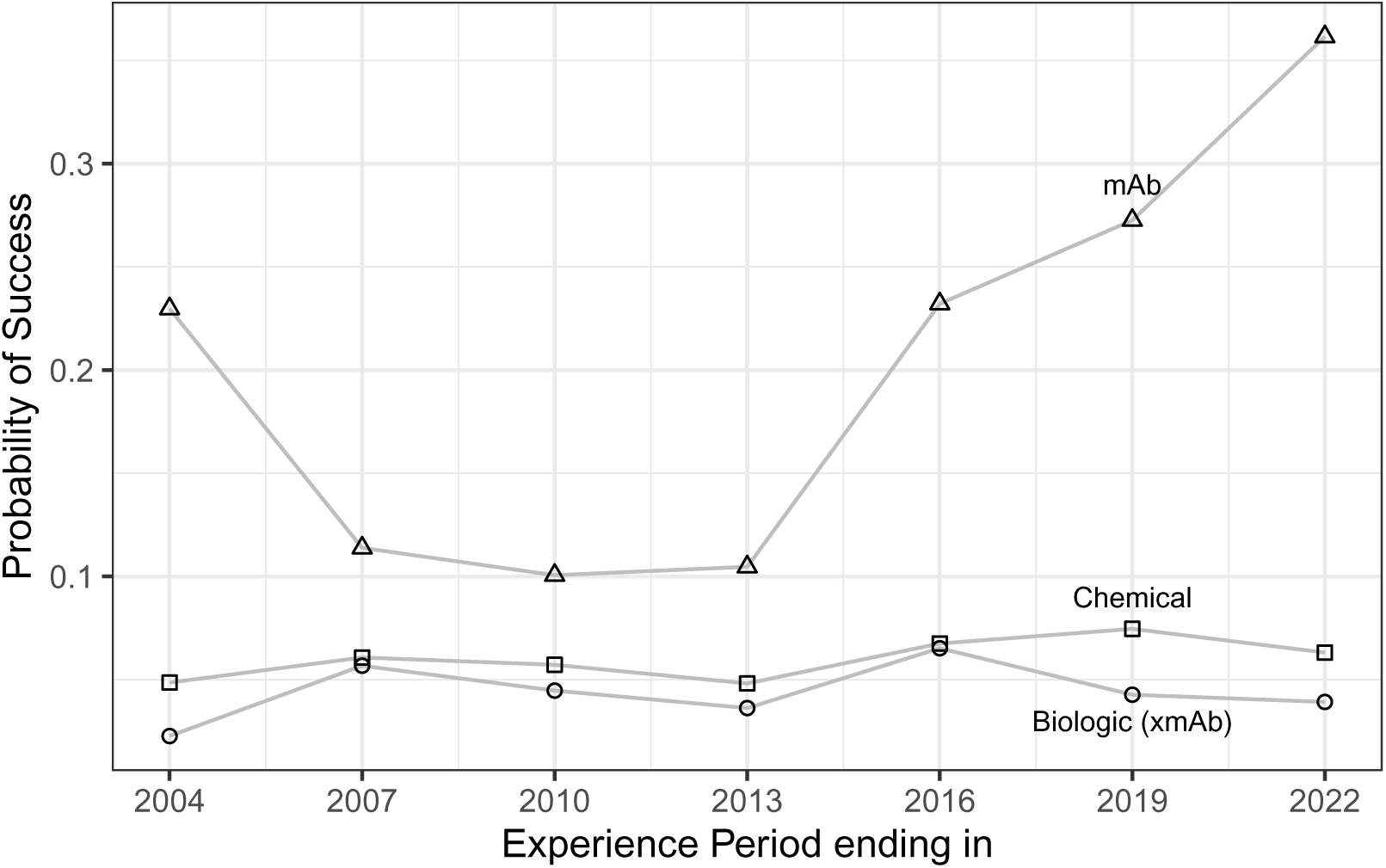
Probabilities of success from Phase I to Approval by experience period and modality. Monoclonal antibody (mAb) drugs have a much larger overall probability of success. The biologic category excludes monoclonal antibody (mAb) drugs.

The probabilities of success by modality are shown in Table 7. Note that for monoclonal antibody drugs the probabilities of approval for the periods from 2005 to 2013 are about 10.7%, but from 2014 to 2019 they jumped up to around 25.3%, and in the last period (2020–2012) the probability of success is 36.2%. In contrast, chemical and biologic (xmAb) drugs have probabilities of approval well below 10%. Chemical drugs’ probability of approval hover around 6.4% during the last 10 years (last 4 rows of Table 7). Biologic (xmAb) drugs have an approximate probability of approval of 4.4% across all periods.

**Table 7.** Overall probabilities of success from Phase I to Approval by experience period and modality. The estimated probability of success (Est.) together with a 95% confidence interval (Low, High) are shown for each modality.

| Period | Monoclonal Antibody |  |  | Chemical |  |  | Biologic (xmAb) |  |  |
| --- | --- | --- | --- | --- | --- | --- | --- | --- | --- |
|  | Low | Est. | Hi | Low | Est. | Hi | Low | Est. | Hi |
| 2002–2004 | 0.085 | 0.230 | 0.563 | 0.039 | 0.049 | 0.085 | 0.008 | 0.023 | 0.188 |
| 2005–2007 | 0.023 | 0.114 | 0.520 | 0.049 | 0.061 | 0.146 | 0.026 | 0.057 | 0.139 |
| 2008–2010 | 0.020 | 0.101 | 0.311 | 0.046 | 0.057 | 0.093 | 0.021 | 0.045 | 0.280 |
| 2011–2013 | 0.047 | 0.105 | 0.273 | 0.039 | 0.048 | 0.096 | 0.019 | 0.036 | 0.094 |
| 2014–2016 | 0.167 | 0.232 | 0.348 | 0.055 | 0.068 | 0.081 | 0.039 | 0.065 | 0.129 |
| 2017–2019 | 0.200 | 0.273 | 0.381 | 0.065 | 0.075 | 0.155 | 0.029 | 0.043 | 0.195 |
| 2020–2022 | 0.301 | 0.362 | 0.433 | 0.054 | 0.063 | 0.077 | 0.027 | 0.039 | 0.118 |

### 3.2. Target

Drugs exert their influence on our bodies by coming into contact with various proteins that are embedded in a cell’s membrane. These cell membrane proteins may be classified as receptors, enzymes, transporters, ion channels, and others. The Pharmaprojects database classifies drugs depending on which of these cell membrane proteins they target; namely, receptor, enzymes, ligand, transporter, ion channel, other, and unspecified. For ligand (*n* = 59), transporter (*n* = 7), and ion channel (*n* = 387) we have very few drugs in the database; thus we have combined them together with the ‘Other’ category. The number of drugs we have in each category are: Receptor (*n* = 6, 537), Enzyme (*n* = 3, 316), Other (*n* = 6, 136), and Unspecified (*n* = 6, 721).

Note that the unspecified category has the largest number of drugs and the lowest overall probabilities of success (from Phase I to Approval). No experience period during the last 20 years has an estimated probability of success above 3.0% for these drugs as shown in Table 8. The remaining categories (receptor, enzyme, and other) have somewhat similar hazard curves shown in Figure 8 and their overall probabilities of success from Phase I to Approval, for each experience period, are similar as displayed in Table 8.

**Figure 8.**
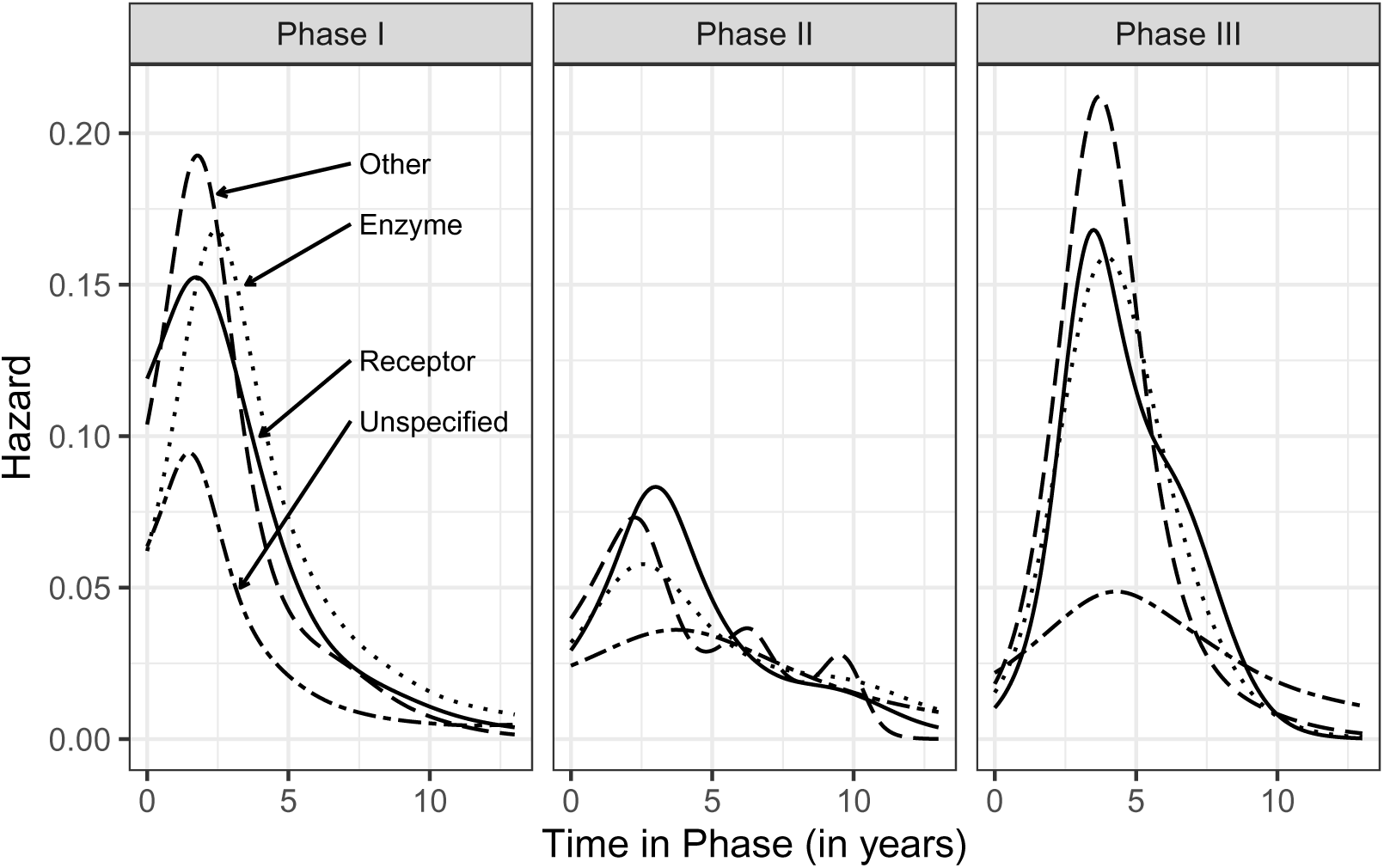
Estimated smooth hazard of transition for the experience period 2017–2019 by development phase and target. The unspecified category has a much lower hazard. The other categories have somewhat similar hazard curves.

**Figure 9.**
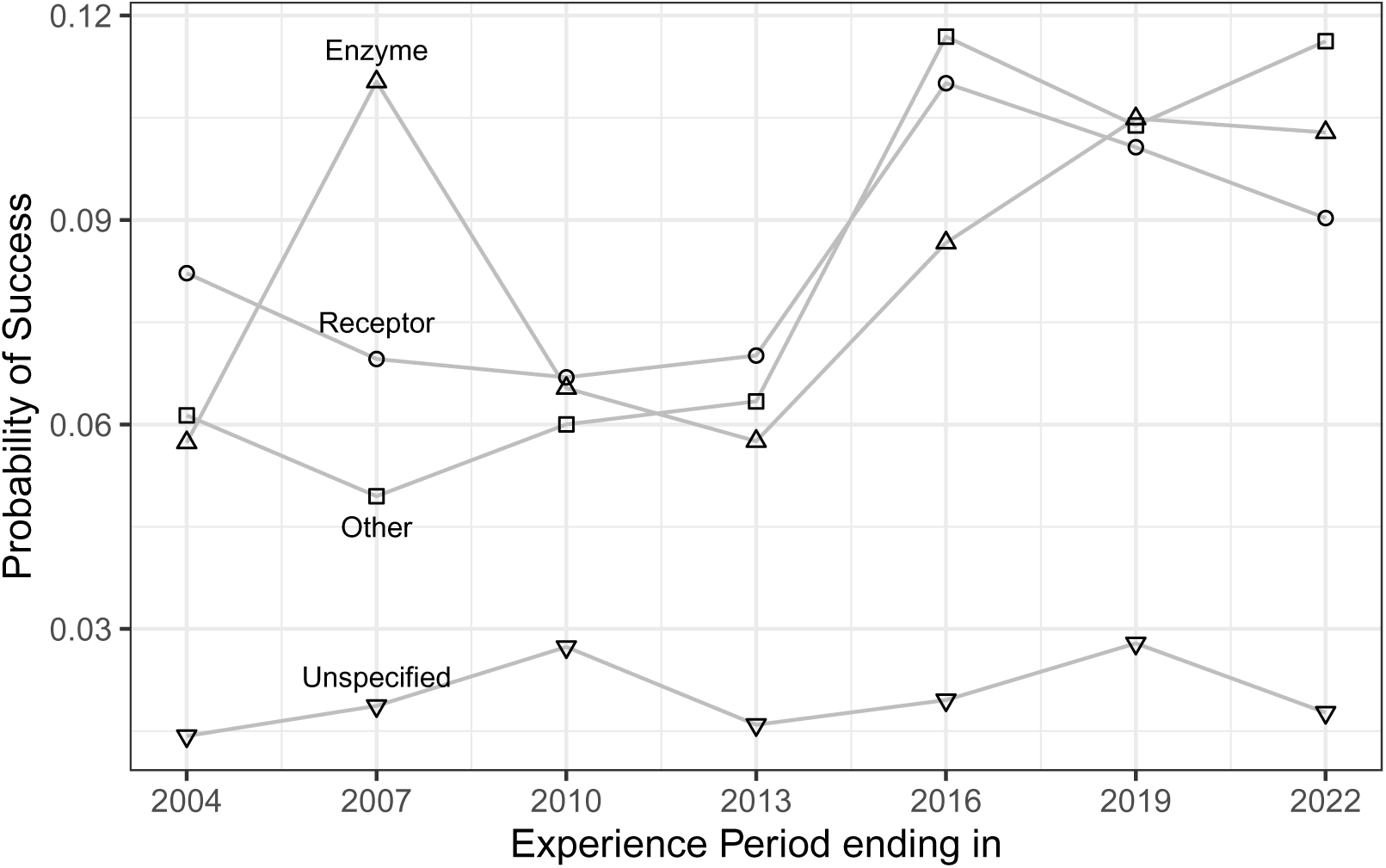
Overall probabilities of success from Phase I to Approval by experience period and target.

**Table 8.** Overall probabilities of success from Phase I to Approval by experience period and target. The estimated probability (Est.) and a lower (Low) and upper (Hi) 95% confidence intervals are shown for every target.

| Period | Enzyme |  |  | Receptor |  |  | Other |  |  | Unspecified |  |  |
| --- | --- | --- | --- | --- | --- | --- | --- | --- | --- | --- | --- | --- |
|  | Low | Est. | Hi | Low | Est. | Hi | Low | Est. | Hi | Low | Est. | Hi |
| 2002–2004 | 0.044 | 0.082 | 0.230 | 0.042 | 0.057 | 0.170 | 0.038 | 0.061 | 0.127 | 0.007 | 0.014 | 0.057 |
| 2005–2007 | 0.038 | 0.070 | 0.138 | 0.077 | 0.110 | 0.178 | 0.032 | 0.049 | 0.136 | 0.008 | 0.019 | 0.100 |
| 2008–2010 | 0.037 | 0.067 | 0.170 | 0.046 | 0.065 | 0.122 | 0.042 | 0.060 | 0.187 | 0.015 | 0.027 | 0.156 |
| 2011–2013 | 0.045 | 0.070 | 0.116 | 0.044 | 0.058 | 0.089 | 0.047 | 0.063 | 0.087 | 0.008 | 0.016 | 0.070 |
| 2014–2016 | 0.076 | 0.110 | 0.227 | 0.067 | 0.087 | 0.184 | 0.088 | 0.117 | 0.157 | 0.011 | 0.020 | 0.036 |
| 2017–2019 | 0.070 | 0.101 | 0.166 | 0.083 | 0.105 | 0.143 | 0.083 | 0.104 | 0.208 | 0.017 | 0.028 | 0.114 |
| 2020–2022 | 0.067 | 0.090 | 0.157 | 0.085 | 0.103 | 0.121 | 0.096 | 0.116 | 0.145 | 0.012 | 0.018 | 0.027 |

### 3.3. Mechanism of Action

Once a drug attaches to a protein on a cell’s membrane there are several ways in which it can influence the cell’s response. An agonist drug will enhance the natural reaction a cell will have whenever this particular receptor is bound. An antagonist drug will block/diminish that natural reaction. An inhibitor drug may suppress a transporter protein from shuttling certain chemicals from inside to outside the cell or vice-versa.

The Pharmaprojects database classifies drugs by the following mechanisms of action: Agonist (*n* = 3, 542), Stimulant (*n* = 2, 058), Antagonist (*n* = 3, 733), Inhibitor (*n* = 8, 118), Other (*n* = 1, 687), and Unidentified (*n* = 3, 572). We have merged the agonist and stimulant drugs into a single category and we have also merged inhibitor and antagonist together. Even though the unidentified category has over three thousand drugs, the number of transitions from Phase III to Approval are extremely small. From calendar years 2002 to 2010 there were no such transitions and in the years 2011 to 2016 we only had single digit number of transitions. The experience period 2020–2022 had only 13 transitions. Therefore, in Figure 10, we do not show the hazard curve for this category and we have also omitted it in Table 9.

**Figure 10.**
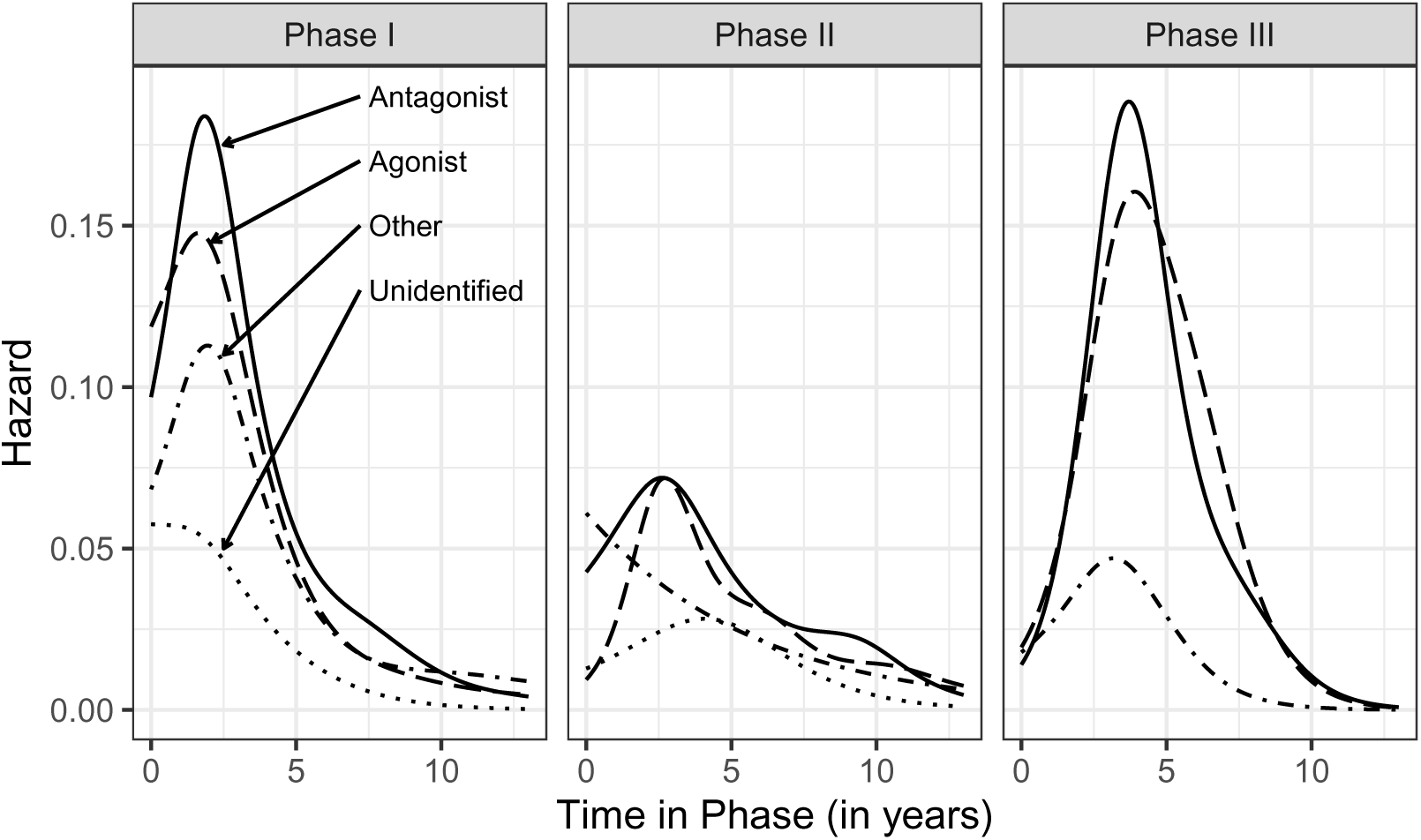
Estimated smooth hazard of transition for experience period 2017–2019 by development phase and mechanism of action.

**Table 9.** Overall probabilities of success from Phase I to Approval by experience period and mechanism of action. The estimated probability (Est.) along with lower (Low) and upper (Hi) points of a 95% confidence interval are shown.

| Period | Agonist |  |  | Antagonist |  |  | Other |  |  |
| --- | --- | --- | --- | --- | --- | --- | --- | --- | --- |
|  | Low | Est. | Hi | Low | Est. | Hi | Low | Est. | Hi |
| 2002–2004 | 0.019 | 0.033 | 0.077 | 0.053 | 0.068 | 0.135 | 0.006 | 0.023 | 0.254 |
| 2005–2007 | 0.044 | 0.068 | 0.144 | 0.060 | 0.077 | 0.154 | 0.005 | 0.028 | 0.144 |
| 2008–2010 | 0.024 | 0.039 | 0.210 | 0.063 | 0.082 | 0.125 | 0.000 | 0.009 | 0.030 |
| 2011–2013 | 0.034 | 0.049 | 0.108 | 0.052 | 0.064 | 0.119 | 0.011 | 0.039 | 0.157 |
| 2014–2016 | 0.053 | 0.076 | 0.154 | 0.090 | 0.107 | 0.126 | 0.027 | 0.060 | 0.208 |
| 2017–2019 | 0.070 | 0.088 | 0.142 | 0.099 | 0.115 | 0.133 | 0.007 | 0.023 | 0.118 |
| 2020–2022 | 0.047 | 0.062 | 0.080 | 0.110 | 0.124 | 0.142 | 0.030 | 0.052 | 0.169 |

As shown in Figure 10 antagonist drugs have a higher hazard compared to the other targets across all three phases leading to higher probabilities of transition and higher probability of approval (Table 9). Also note that during the period 2011 to 2022, antagonist drugs show an increasing probability of success from Phase I to approval. For agonist drugs, we also see the increasing pattern in the probabilities of success except for the last period (2020–2022) where we have a decline. Figure 11 shows a steady increasing calendar year trend across all three mechanisms of action.

**Figure 11.**
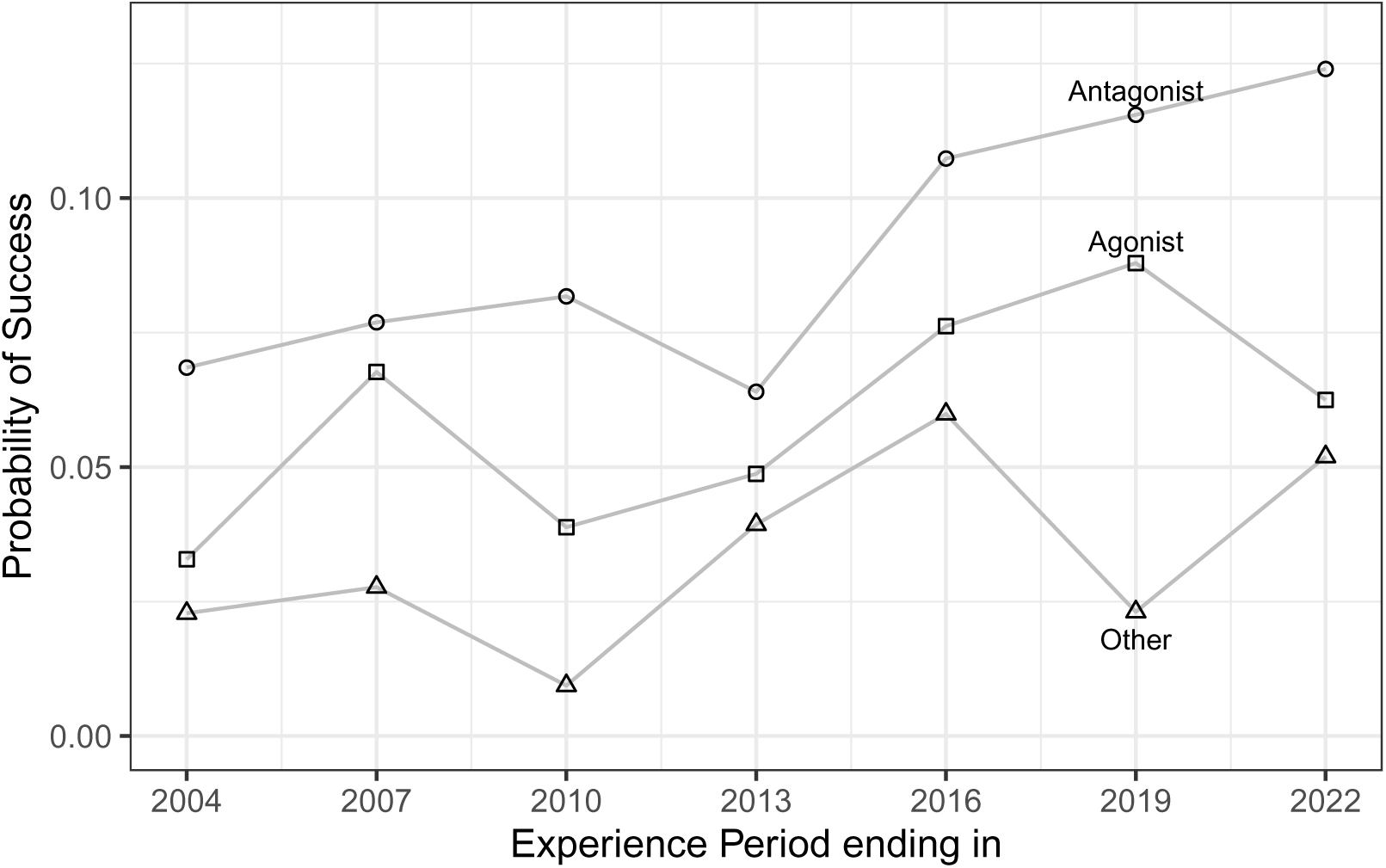
Overall probability of success from Phase I to Approval by mechanism of action and experience period. Note that the ‘Other’ mechanism of action is always below the others and that all four of them show a positive trend as calendar time increases.

### 3.4. Therapeutic Indication

Another characteristic available in the Pharmaprojects database relates to the indication the drug is intended for. We have 16 different indications ranging from ‘Alimentary/Metabolic’ to ‘Unspecified’ and Table 10 gives the complete list. A drug may have several indications and thus these categories do not partition our data set into mutually exclusive subsets. But around 76% of the drugs have only one indication. Therefore, we have chosen to use the first named indication as *the* indication for the drug.

**Table 10.** List of therapeutic indication groups.

| Indication | Indication | Indication |
| --- | --- | --- |
| Anti-cancer | Dermatological | Respiratory |
| Anti-infective | Genitourinary (incl. sex hormones) | Sensory |
| Anti-parasitic | Hormonal (excl. sex hormones) | Miscellaneous |
| Alimentary/Metabolic | Immunological | Unspecified |
| Blood & Clotting | Musculoskeletal |  |
| Cardiovascular | Neurological |  |

Figure 12 displays the hazard of transition for the top 4 indications during experience the period 2017–2019. Note that for all three clinical trial phases, ‘Alimentary/Metabolic’ and ‘Anti-infective’drugs have, in general, the highest hazard curves and so these drugs will have higher probabilities of success. On the other hand, ‘Neurological’ drugs have the lowest hazard curves across all three phases.

**Figure 12.**
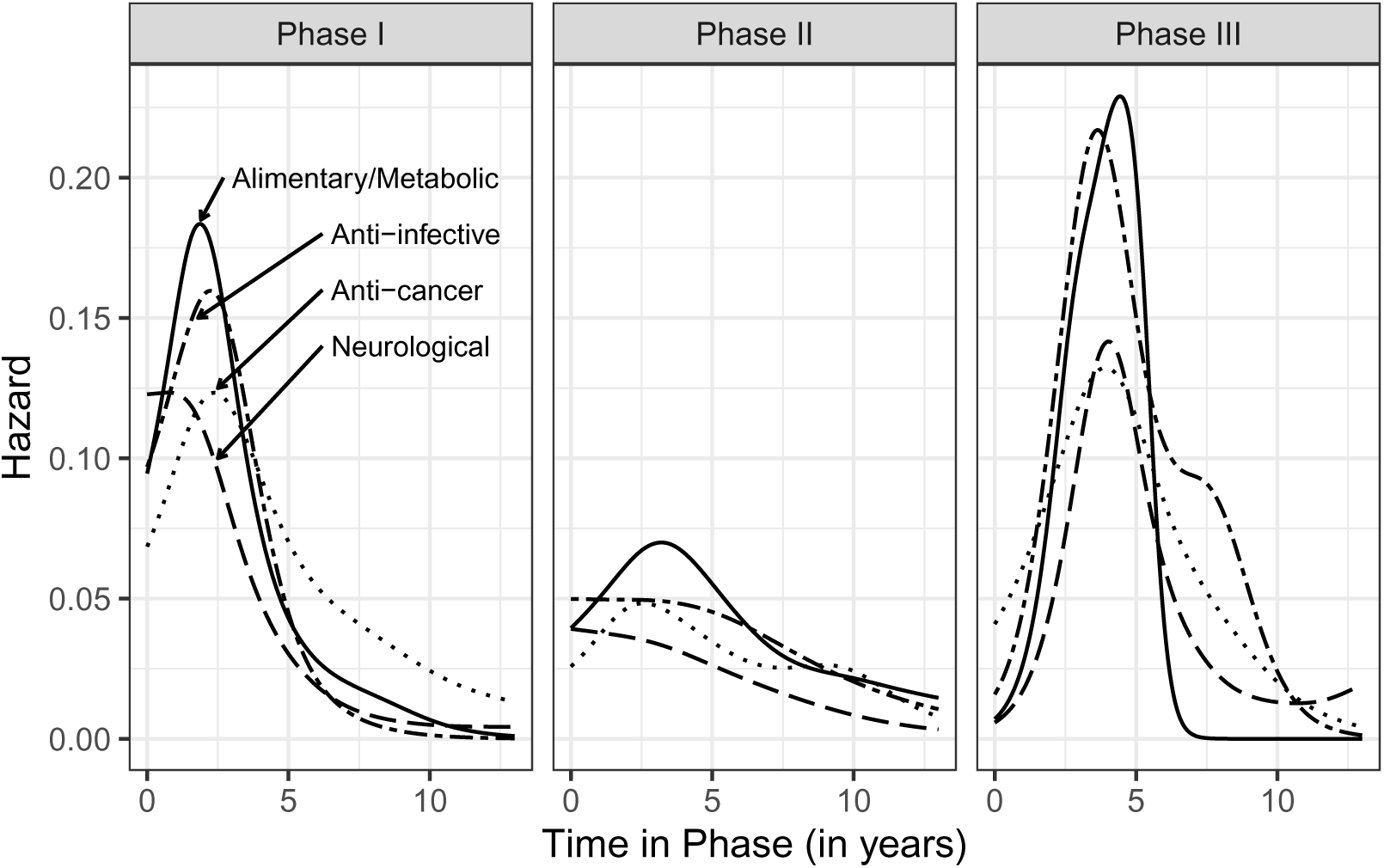
Estimated smooth hazard of transition for experience period 2017–2019 by development phase and indication for the top 4 indication categories.

The hazard curves for anti-cancer drugs show some interesting patterns. For Phase I, we see a very thick tail. After five years the hazard curve for these drugs, the dotted curve in Figure 12, is much higher than the rest and it continues all the way to the end of the time axis (*t* = 13). This suggests that anti-cancer drugs take a longer time to transition out of Phase I. During Phase II we see that the hazard curve for these drugs has two peaks: the first one around at 2.5 years and the second one around 9 years. One possibility for this pattern is that we have two classes of anti-cancer drugs. One class would have short development time and the other class a longer one. Another possibility is that some anti-cancer drugs, after failing at their original indication, may be re-purposed and successfully transition out of the phase at a later date.

Table 11 displays the probabilities of success from Phase I to Approval by experience period for the top four indications along with lower and upper 95% confidence intervals. Anti-infective and Alimentary/Metabolic drugs have a probability of success that is fairly stable around 10.6%.

**Table 11.** Overall probabilities of success from Phase I to Approval by experience period for the top four indications. The estimated probability (Est.) along with lower (Low) and upper (Hi) points of a 95% confidence intervals are shown.

| Period | Anti-Infective |  |  | Alimentary/Metabolic |  |  | Anti-Cancer |  |  | Neurological |  |  |
| --- | --- | --- | --- | --- | --- | --- | --- | --- | --- | --- | --- | --- |
|  | Low | Est. | Hi | Low | Est. | Hi | Low | Est. | Hi | Low | Est. | Hi |
| 2002–2004 | 0.070 | 0.118 | 0.316 | 0.062 | 0.099 | 0.448 | 0.025 | 0.049 | 0.190 | 0.010 | 0.026 | 0.113 |
| 2005–2007 | 0.046 | 0.080 | 0.196 | 0.077 | 0.120 | 0.350 | 0.020 | 0.042 | 0.077 | 0.025 | 0.047 | 0.134 |
| 2008–2010 | 0.038 | 0.078 | 0.209 | 0.056 | 0.100 | 0.222 | 0.033 | 0.058 | 0.199 | 0.013 | 0.027 | 0.085 |
| 2011–2013 | 0.063 | 0.112 | 0.226 | 0.045 | 0.066 | 0.115 | 0.039 | 0.062 | 0.271 | 0.010 | 0.020 | 0.071 |
| 2014–2016 | 0.060 | 0.097 | 0.223 | 0.088 | 0.120 | 0.210 | 0.041 | 0.063 | 0.130 | 0.036 | 0.060 | 0.129 |
| 2017–2019 | 0.084 | 0.117 | 0.220 | 0.086 | 0.110 | 0.148 | 0.071 | 0.095 | 0.123 | 0.027 | 0.043 | 0.125 |
| 2020–2022 | 0.045 | 0.066 | 0.116 | 0.096 | 0.125 | 0.254 | 0.061 | 0.078 | 0.104 | 0.037 | 0.054 | 0.106 |

Anti-cancer drugs have an increasing trend shown in Figure 13 even though the last period dipped below the prior estimate. Neurological drugs have the lowest probabilities of success across all experience periods, and like anti-cancer drugs, there is a positive calendar year trend.

**Figure 13.**
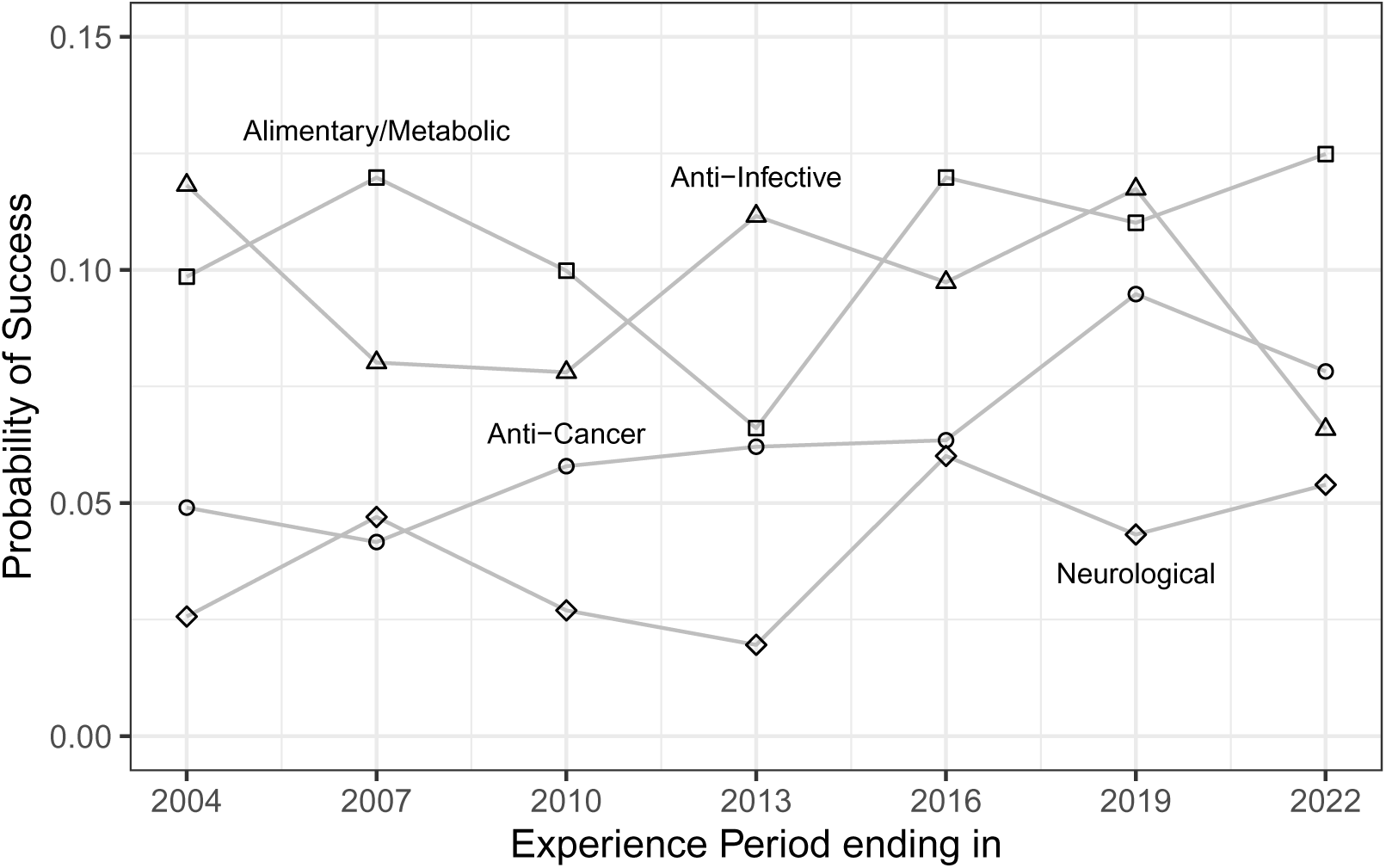
Overall probability of success from Phase I to Approval by indication and experience period.

## 4. Application

In this section, we provide a simple application evaluating various probabilities for a portfolio of drugs. Consider a company that has four drugs in various stages of development. Let the drugs be A, B, C, and D and assume that drug A has been in a Phase I clinical trial for 2 quarters.

Drugs B and C have been in Phase II clinical trials for 4 and 6 quarters, respectively. And finally, assume that drug D has been in a Phase III clinical trial for 5 quarters.

What is the probability that at least one of these drugs will transition to the next phase within the next 2, 4, or 6 quarters?

To answer this question we assume that our work estimating the hazard and survival functions from the experience period 2020–2022 is appropriate. Table 12 shows only the entries of the survival function necessary to evaluate this portfolio and Table 13 gives the results.

**Table 12.** Smooth survival probabilities estimated from experience period 2020–2022 across all drugs for each phase of human clinical trials.

| Quarter | Time in<br>Phase (yrs) | Survival Probabilities for |  |  |
| --- | --- | --- | --- | --- |
|  |  | Phase I | Phase II | Phase III |
| 0 | 0.00 | 1.00000 | 1.00000 | 1.00000 |
| 1 | 0.25 | 0.98224 | 0.99016 | 0.99158 |
| 2 | 0.50 | 0.96246 | 0.97984 | 0.98240 |
| 3 | 0.75 | 0.94051 | 0.96902 | 0.97240 |
| 4 | 1.00 | 0.91630 | 0.95768 | 0.96149 |
| 5 | 1.25 | 0.89000 | 0.94586 | 0.94957 |
| 6 | 1.50 | 0.86202 | 0.93362 | 0.93649 |
| 7 | 1.75 | 0.83309 | 0.92103 | 0.92205 |
| 8 | 2.00 | 0.80411 | 0.90823 | 0.90604 |
| 9 | 2.25 | 0.77594 | 0.89537 | 0.88833 |
| 10 | 2.50 | 0.74937 | 0.88265 | 0.86890 |
| 11 | 2.75 | 0.72501 | 0.87027 | 0.84789 |
| 12 | 3.00 | 0.70319 | 0.85839 | 0.82559 |

**Table 13.** Probabilities of transition within *n* quarters given that a drug has been in a particular phase for some given number of quarters.

| Drug | Phase | Time Spent in<br>Phase (Q's) | Probability (in %)<br>of Transition within |  |  |
| --- | --- | --- | --- | --- | --- |
|  |  |  | 2Qs | 4Qs | 6Qs |
| A | I | 2 | 4.796 | 10.437 | 16.453 |
| B | II | 4 | 2.513 | 5.164 | 7.835 |
| C | II | 6 | 2.719 | 5.459 | 8.057 |
| D | III | 5 | 2.899 | 6.450 | 10.708 |

For example, the probability that drug B will transition within the next 4 quarters is calculated as follows

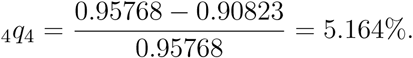

The numerator represents the fraction of drugs that transitioned between quarter 4 and quarter 8 and the denominator represents the fraction of drugs that are still available to transition at quarter 4.

Thus, using the above probabilities we can calculate the chances of *at least one* drug transitioning within the next two quarters as

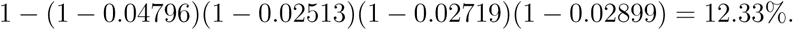

Similarly, the probability of at least one drug transitioning in the next 4 and 6 quarters would be 24.88% and 36.78%, respectively.

The choice of a quarter as a unit of time is arbitrary and we could easily change it to yearly or any other time span.

## 5. Concluding Remarks

The development of a compound into a drug that humans can use to manage/cure a disease is a laborious and risky endeavor. Less then 1 in 10 drug candidates that begin human clinical trials are approved for human use. But the probabilities of success vary significantly across different drug characteristics, such as, modality, mechanism of action, and indication.

This study was based on a large industry data base, Pharmaprojects, were we have taken a cross-sectional approach to estimating the probability that a drug will transition successfully out of a phase of development. This industry data base has some limitations: the information it contains is based on what drug sponsors are willing to share with the public, and the starting dates of clinical trials do not have the corresponding ending dates. Lack of ending dates for clinical trials required us to use a proxy and thus our estimation of the length of time spent in a given phase of development is slightly overstated.

Our approach differs from the traditional longitudinal study in that we are estimating the probabilities of transition experienced during a calendar period of observation and thus we are able to track the dynamics of the process as it unfolds across time. With a longitudinal study we would have to observe these drugs for many years before we could have a reliable estimate of the probabilities of transition.

The probabilities we have estimated do not correspond to a particular cohort of drugs. But if we assume that the development process is stable, we can use them to project the transitions we will see going forward. This is a similar assumption that we would have to make had we done a longitudinal study and used it to project outcomes for future cohorts.

For each calendar period of observation, we summarized the transition experience from a clinical trial phase through a life table, obtained raw estimates of the transition hazard, and graduated them via a generalized additive model with a Poisson distribution and linear predictor with a smooth on the time spent inside a phase. Based on the smooth hazard rates, we calculated survival and cumulative incidence curves as well as conditional probabilities of transition.

While the overall probability of success for a drug candidate that enters the human clinical trials is slightly below 10%, there are significant differences in this rate depending on broad characteristics of the drugs in question. For example, we showed that monoclonal antibody drugs have a much higher probability of success, particularly after the mid 2010’s. Similarly, drugs that suppress the natural reaction of cell, namely antagonist drugs, have a higher rate of success. But more importantly, across target, mechanism of action, and certain indications (anti-cancer and neurological) we see an upward trend in their probabilities of success (see Figure 9, Figure 11, and Figure 13). This suggests that the development process is improving, albeit slowly, and thus in the coming years we should see a higher rate of drug approvals. Also based on our work, we can evaluate various probabilistic statements for a portfolio of drugs.

## Data Availability

The data used in this study is commercially available from PharmaProjects (Citeline) and available upon reasonable request to the authors.

https://www.citeline.com/en/products-services/clinical/pharmaprojects

## Acknowledgments & Funding

This work was supported by a grant from the National Biomedical Research Foundation to Bentley University.

